# Multimodal diagnosis of Alzheimer’s disease through causal imaging markers and risk factors

**DOI:** 10.64898/2026.05.01.26352207

**Authors:** Geetha Chilla

**Author notes:** 8 Biomedical Grove, Neuros 05-01, Singapore, 138665.

## Abstract

**Objectives:** Stage-sensitive markers can aid in early diagnosis of Alzheimer’s disease (AD) and can improve sensitivity, performance and interpretability. In this study, causal markers from longitudinal imaging data were extracted and integrated with risk factors to improve diagnostic models.

**Data Description:** OASIS-3, a longitudinal dataset consisting of 613 controls and 214 cases with very mild to moderate Alzheimer’s disease is used for this study. A meta model was built using a predisposition model built from risk factors, a stage-sensitization model built from MRI markers at various stages of atrophy and a confirmatory model built using PET markers. The meta model achieved good diagnostic performance (accuracy = 93%, sensitivity = 80%, specificity = 95%). Exclusion of PET data achieved comparable performance (accuracy = 91%, sensitivity = 85%, specificity = 92%). The results demonstrate that integrating causal pathological markers with risk factors improves diagnosis and aids in elucidating stage-specific patterns of AD.

## 1. Objective

Alzheimer’s disease (AD), characterized by buildup of amyloid plaques and neurofibrillary tau tangles in the brain is the most common cause of dementia worldwide, contributing to 60-80% of dementia cases in individuals aged 65 and above. Current diagnosis is based on clinical examination, laboratory tests, neuropsychological assessments and imaging if required. These tests are used to assess cognitive decline, functional impairment and evaluate AD-specific changes such as amyloid-β plaques & tau tangles accumulation and hippocampal atrophy [1]. Severity can be estimated clinically using Clinical Dementia Rating (CDR) which evaluates cognition and function in 6 domains, namely memory, orientation, judgment and problem-solving, community affairs, home and hobbies, and personal care. This score ranges from 0 (no cognitive impairment) to 3 (severe cognitive impairment), with 0.5 indicating questionable impairment, 1 indicating mild cognitive impairment (MCI) and 2 indicating moderate cognitive impairment.

However, definitive diagnosis and corresponding staging for AD is only through post-mortem histopathological examination and determination of amyloid and tau buildup in brain tissue, using mechanisms such as Braak staging. This staging system classifies progression of AD into six stages, reflecting the spread of neurofibrillary tangles (NFT) in brain [2, 3]. Stages I-II, involve NFT deposition in transentorhinal and entorhinal regions. However, at these stages, cognition is often normal with typical CDR scale of 0. In stages III-IV (Limbic stages), NFTs spread to the hippocampus and other limbic structures, potentially accompanied by mild cognitive symptoms with a typical CDR score of 0.5. Stages V-VI, referred to as Neocortical stages, feature extensive NFTs in neocortical areas, associated with severe cognitive decline and clinical dementia (CDR ≥ 1) [4–6].

Attempts to find clinical in-vivo and in-vitro markers that can act as proxy for such histopathology diagnosis and staging are ongoing, either targeted towards identifying core AD processes such as amyloid-β proteinopathy and tau pathology or non-AD specific processes such as neurodegeneration [7]. PET imaging is suitable for in-vivo evaluation of amyloid-β and tau burden, which in turn has been shown to have positive correlation with cognitive decline [8–10]. New tracers are being studied to improve specificity, sensitivity and reducing off-target binding [11]. Additionally, structural MR imaging markers are also being studied to bridge the gap between clinical and biological progression. Specifically, structural MRI was used to estimate NFT burden where significant associations of structural atrophy with Braak stages in several studies [12, 13]. Dallaire-Théroux *et al*. [12] identified 59 regions which could discriminate between transentorhinal, limbic, and isocortical Braak NFT stages, validated using postmortem data, with 62.4% accuracy. Similarly, several MR measures such as hippocampal atrophy and cortical thickness have also been studied and correlated with CDR scores [14–16].

In addition to imaging, biological, clinical and socio-economic risk factors of AD are also been investigated. Apolipoprotein E (APOE) ε4 allele was linked to increased AD risk and faster progression while ε2 is considered to have protective effect [17–19]. AD prevalence was also found to be more in women and is attributed to biological and sex-specific differences concerning hormonal changes and pregnancy [20]. Parental, especially maternal family history of dementia has been positively associated with AD and so is the presence of certain medical conditions in the patient such as cardiovascular diseases, obesity and depression [21, 22]. While these are not confirmatory factors, they increase AD risk of an individual at later stages of life.

In an effort to better understand the disease and its progression as well as for accurate diagnosis multimodal approaches are increasingly being considered. In [23], authors used features from amyloid PET, tau PET and MRI to classify healthy controls, MCI and AD patients. They found that tau-PET to be the most effective modality for classification compared to other imaging types but noted that multimodal approach delivers overall improved performance. However, they did not find increased performance with the inclusion of risk factors such as age, gender, APOE ε4 and education. Similar multimodal classification was carried out in studies [24, 25] using MRI atrophy and PET data to classify normal, MCI and AD patients with decent predictive accuracy. Deep learning based multimodal approaches have been carried out in other studies [26, 27]. However, the main drawback of these studies is lack of features sensitive to AD-specific stages which affect interpretability of these features in disease context. Efforts to model features sensitive to AD-specific pathways are ongoing [28] in population sub-groups.

In line with this and in an effort to make diagnostic models sensitive to AD-specific progression, this study is carried out where imaging markers were extracted using Bayesian models that are associated with disease and/or atrophy levels from longitudinal data. Combining these with key risk factors, such as age, sex, genetic phenotyping, parental and subject medical history, an ensemble model was built using for AD likelihood prediction. This approach of extracting causal factors prior to integration combines strengths of multimodality approach and helps in improving interpretability by reducing model complexity enabling easier deployment to clinics [29].

## 2. Data Description

### 2.1 Data and preparation

Longitudinal data from OASIS-3 [30], consisting of data from cognitively normal individuals and mild-moderate dementia (CDR 0 – 2) patients, aged 42-95 years, is used for this study. This dataset consists of longitudinal neuroimaging from MRI and PET (for a subset), clinical and demographic information from APOE subtyping, family history of dementia, patient medical history and cognitive information from MMSE and CDR scores and clinician coded dementia diagnosis.

Using clinician coded dementia diagnosis (dx1-dx5) available in the *ADRC Clinical Data*, data from subjects with non-AD type dementia such as DLBD, FTD, vascular dementia, incipient non-AD dementia and other causes such as head trauma and secondary neurological diagnosis like Parkinson’s disease, hydrocephalus has been excluded. Data is then split into (a) healthy controls (cognitively normal group), (b) transition group (cognitively normal at first timepoint and diagnosed with AD at later timepoints) and (c) AD group (diagnosed with AD since first visit) which formed basis for AD stage categorization in the context of this study. For all the subjects, MRI derived morphometry measures namely, subcortical & cortical volumes (n = 97) and cortical thickness (n = 68) were extracted. After removing any outliers using Mahalanobis distance, the dataset samples were 827 subjects - 613 from cognitively normal individuals, 56 from transition group and 158 from AD group.

#### 2.1.1 Volumetric data correction and normalization

Inter-subject volumetric variability corresponding to intracranial volume (ICV) was corrected using analysis of covariance method, where *V*_*corr*_ is a ICV adjusted volume and b is the slope of regression of raw volume *V*_*raw*_ with ICV [31].

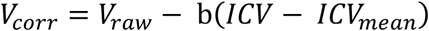

Following this, a log transformation was carried out on these ICV corrected volumes for normalization of morphometry data.

#### 2.1.2 AD subgroups categorization

158 subjects’ data of AD group is further clustered into subgroups based on volumetric atrophy and cortical thinning levels, using PCA and hierarchical clustering. 62 principal components, explaining 95% variation of the data, were obtained from 165 thickness and ICV-adjusted volume measures. These principal components were used to carry out hierarchical clustering, which revealed 3 clusters at increasing levels of volumetric atrophy and cortical thinning, hereafter referred to as early AD, intermediate AD and advanced AD groups for the remainder of this study.

Corresponding MMSE and CDR, along with its subscales, have been merged with this data based on the closest timepoint such that each neuroimaging data has one set of associated CDR and MMSE data for all individuals at all time points. An overview of the data pre-processing is given in Fig. 1.

**Fig. 1.**
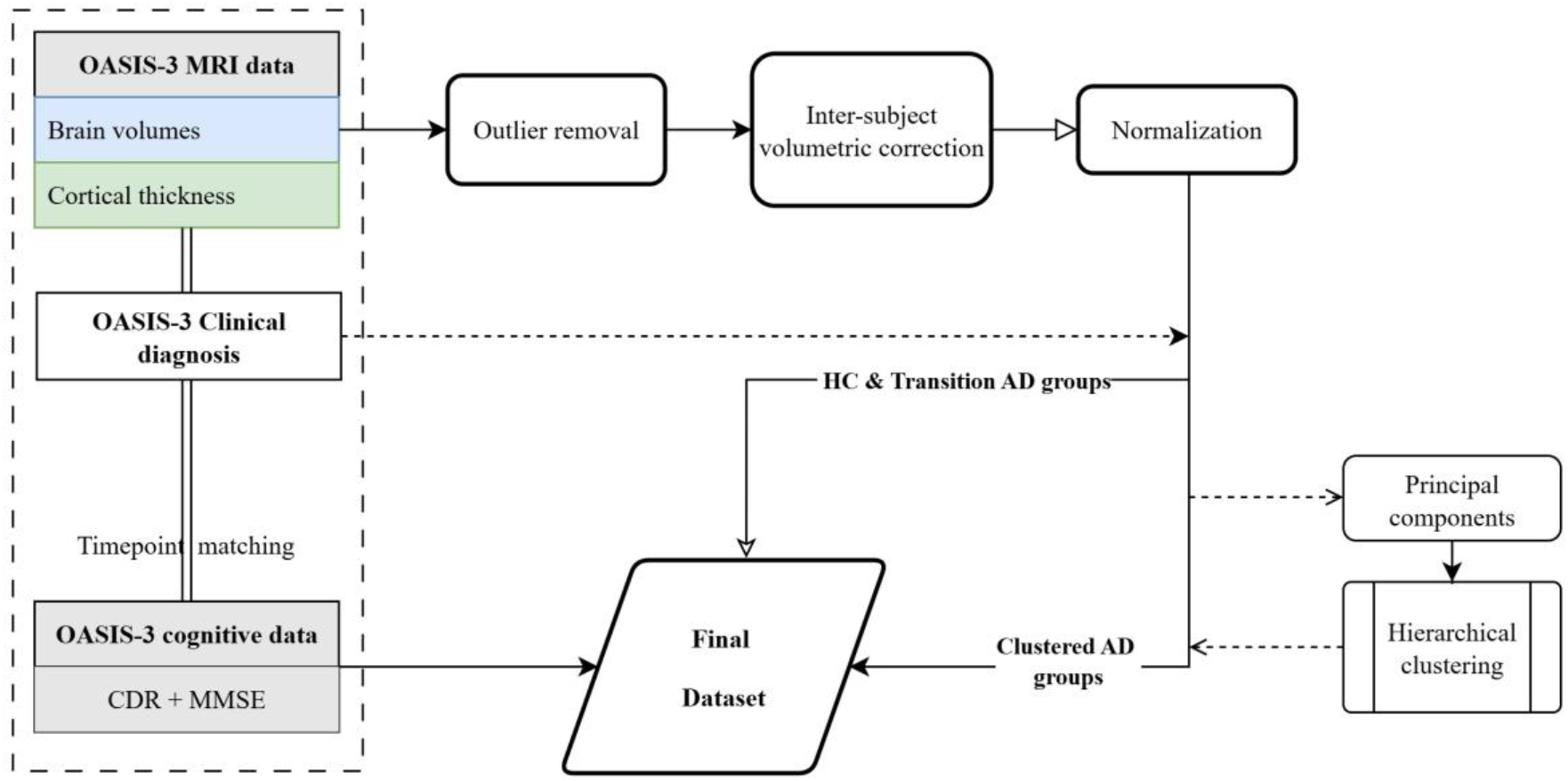
Overview of OASIS-3 data pre-processing

Although these AD groups have been clustered based on atrophy alone, they correlated well with age and MMSE scores, with early AD group being the youngest and having highest mean MMSE scores and advanced AD group being the oldest and having lowest MMSE scores. Additionally, the proportion of patients with moderate cognitive impairment was 7.4% in advanced AD group, from 0% in transition/early/ intermediate AD groups, suggesting the utility of brain atrophy levels for clinical cognitive and functional impairment correlations. Key characteristics of the final dataset from these five categories, healthy controls (HC), and four AD groups is given in Table 1. Proportion of these groups in severity of CDR subscales is given in Fig. 2.

**Table 1.** Characteristics of final processed dataset.

| **OASIS-3 data** | **Healthy Controls** | **Transition AD** | **Early AD** | **Intermediate AD** | **Advanced AD** |
| --- | --- | --- | --- | --- | --- |
| **Subjects**<br>(n=829) | 613 | 56 | 37 | 73 | 48 |
| **Datapoints** | 1514 | 105 | 45 | 83 | 54 |
| **Age, years** | 69.04±9.25 | 76.06±7.01 | 71.28±7.31 | 76.23±7.64 | 78.21±6.89 |
| **Sex** |  |  |  |  |  |
| Female | 380 | 25 | 18 | 37 | 24 |
| Male | 233 | 31 | 19 | 36 | 24 |
| **MMSE score** | 29.22±1.05 | 27.76±2.21 | 25.8±3.42 | 24.92±3.2 | 22.2±4.39 |
| **CDR, score=, %** |  |  |  |  |  |
| 0 | 100 | 34.3 | 0 | 0 | 0 |
| 0.5 | 0 | 63.8 | 64.4 | 75.9 | 33.3 |
| 1 | 0 | 1.9 | 35.6 | 24.1 | 59.3 |
| 2 | 0 | 0 | 0 | 0 | 7.4 |

**Fig. 2.**
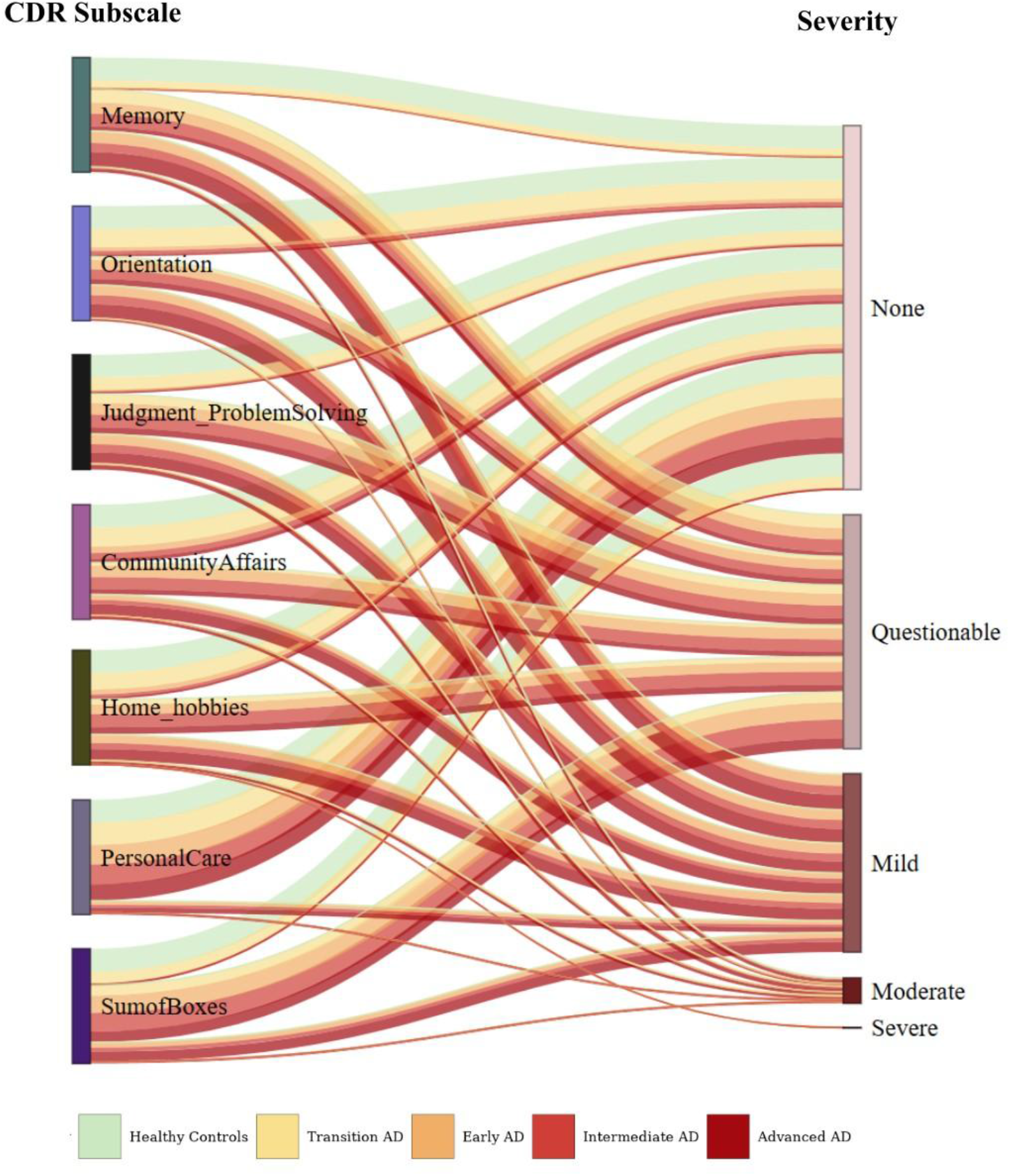
Distribution of severity levels (%) across CDR subscales in HC and AD groups

#### 2.1.3 PET data

PET imaging, carried out using two tracers – [11C]-Pittsburg Compound B (PiB) and [18F]-Florbetapir (AV45), was available for a subset of data (AV45 & PiB: HC – 423 & 801, Transition AD group −22 & 30, AD group – 26 & 35). The data was registered with MRI, Freesurfer based regional measurements were extracted and corrected for partial volume effects. 166 features including global amyloid deposition, quantified on the Centiloid scale, and regional standard uptake value ratios (SUVR) in left and right hemispheres, were used in this study. Given the smaller sample size, AD cohort has been analyzed as one group instead of early, intermediate and advanced groups, unlike MRI data.

#### 2.1.4 Non-imaging clinical data

Additional data (APOE genotyping, family history and patient medical history) was also available for a subset of these subjects, which was matched to MRI and PET data based on the closest timepoint.

### 2.2 Feature extraction

#### 2.2.1 Imaging data

Given the longitudinal nature of the data and limited timepoints, feature selection has been carried out using Bayesian networks and generalized linear mixed modeling (GLMM) approach through MXM [32] package of R. Specifically, Max-Min Parents and Children (MMPC) GLMM [33] is employed, which identifies variables that form Markov blanket of the target (HC/AD) in the Bayesian network using a forward-backward filter approach, while accounting for within-subject correlation. Multiple conditional independence tests are performed in MMPC where key variables are selected using log-likelihood tests that result in a minimal but highly predictive feature subset.

#### 2.2.2 Non-imaging data

Given the lower sample size as well as lack of consistent sample sizes across different items on clinical and demographic data, univariate analyses were carried on individual item to evaluate its association with the disease and without any data imputation. Binary logistic regression models were used to extract factors strongly associated with the disease, except for modelling age where generalized linear additive mixed model was used.

### 2.3 Multimodal diagnostic system

Using features extracted from MRI, PET and assessed risk factors, a multimodal diagnostic system built on three independent binomial logistic regression models was developed.

The first model is a predisposition model built using features identified from cross-sectional clinical and demographic data whose prediction probabilities indicate individuals’ risk of predisposition towards AD. While significant features were selected using univariate analyses, the model, however, was built using a subset of data with significant features, keeping rows with <50% missing data, and missing values then imputed using the MICE (Multiple Imputation by Chained Equations) method.

The second model is a stage sensitization model built using key brain atrophy features of early, intermediate and advanced AD stages identified from MRI. Given that these markers are indicative of progressive atrophy levels in AD, this model has the potential to identify patients at earlier stages of AD.

The third model is an amyloid proxy model based on PET imaging which is sensitive towards amyloid deposition in brain. The extracted features represent key brain areas where tracer uptake can distinguish between healthy individuals and AD patients.

Using the disease probabilities obtained from these predisposition, stage-sensitization and amyloid proxy models, a metamodel was built using xgboost to classify healthy individuals and stage-agnostic AD patients.

## 3. Results

### 3.1 Non-imaging clinical markers and predisposition model

#### Age

Age was significantly and non-linearly associated with AD when evaluated using generalized additive mixed models, exhibiting a risk of 50% after an age of 86.3 years, as shown in Fig. 3A.

**Fig. 3.**
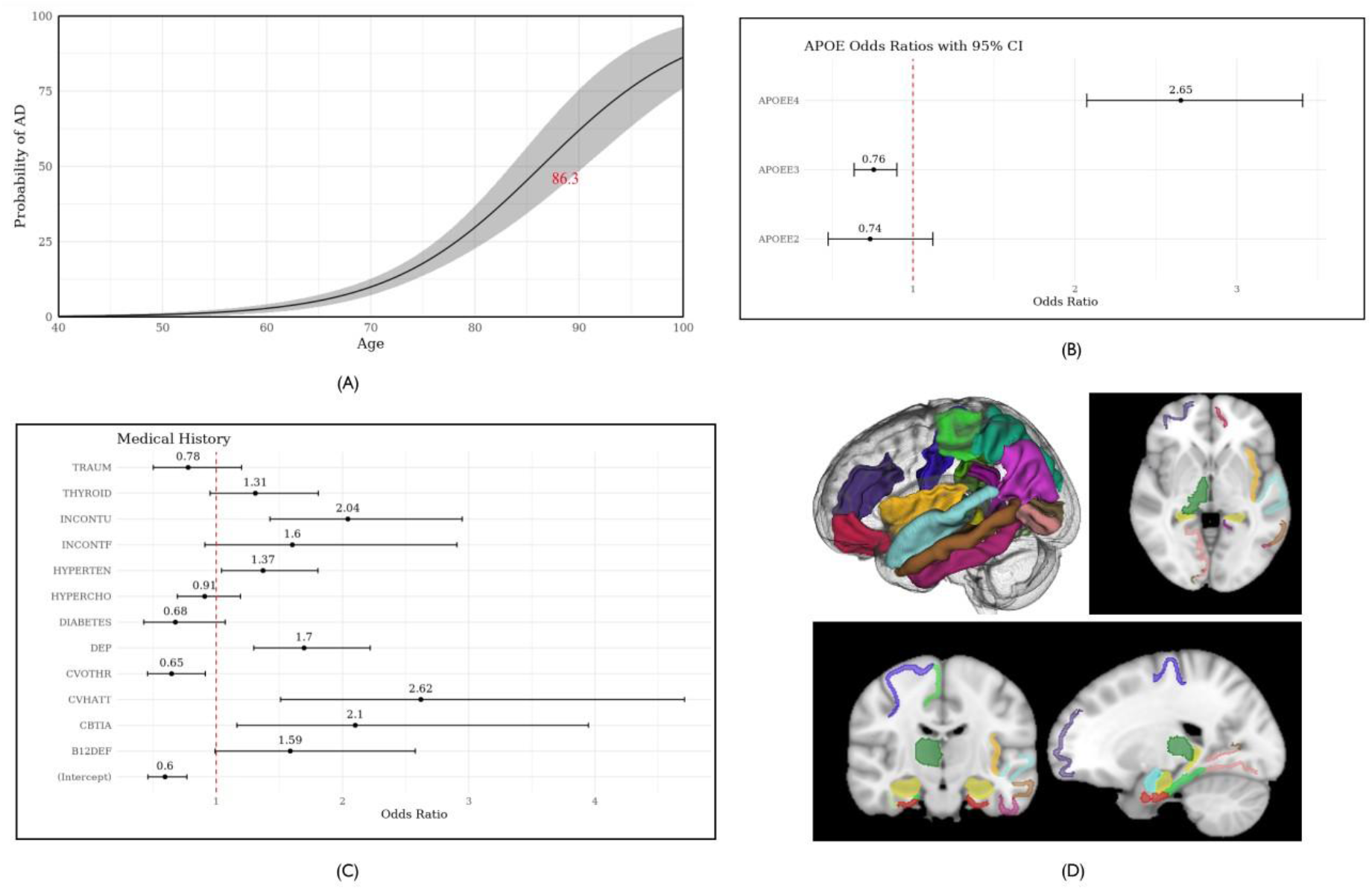
AD risk profiles associated with (A) age showing 50% risk at 86.3 years, (B) APOE status, with Odds-Ratios and 95% CI, and (C) medical conditions, with Odds-Ratios and 95% CI. (D) shows MRI regions indicative of AD at various stages of atrophy stages.

#### APOE status

While specific genotyping did not reveal any significant associations with the disease, it was found that E2 carriers (22 and 23) were associated with lower risk and E4 carriers (24, 34 and 44) with higher risk of AD compared to homozygous E3 genotype (33). For this study, e2/e4 genotype was considered as E4 carriers, consistent with Goldberg *et al*. [34] who found that e2/e4 genotype has similar risk profile for AD pathology as E4 carriers. However, membership of e2/e4 into E2 carriers did not change the findings in study. Risk profile of these carriers is given in Fig. 3B.

#### Socio-demographic factors and parental history

No significant correlation between AD and sex, handedness or paternal history of dementia was found. However, maternal dementia was positively associated with likelihood of AD.

#### Medical history

A total of 14 conditions including depression, hypertension, hypercholesterolemia, diabetes, B12 deficiency, thyroid diseases, urinary/fecal incontinence, alcohol abuse, any cardiovascular diseases including suffering from cardiac arrest, atrial fibrillation, having any cardiovascular procedures such as angioplasty, cardiac bypass or pacemaker procedures carried out, cerebrovascular diseases or conditions including traumatic brain injury, transient ischemic attack were tested. Additionally, interaction with APOE carrier status on risk of AD was also tested. Of the tested conditions, significant link was found between AD and cardiac arrest, transient ischemic attack, depression, hypertension, b12 deficiency, thyroid diseases, fecal and urinary incontinence. In addition to these conditions, cerebrovascular diseases, traumatic injury, hypercholesterolemia and diabetes for specific APOE carrier types also seem to increase risk of AD. Risk profiles Odds for these are given in Fig. 3C.

#### 3.1.1. Predisposition model

On test dataset, logistic regression model built using these non-imaging markers demonstrated an accuracy of 69%, sensitivity and specificity of 70% and 68% with an AUC of 0.76.

### 3.2 MRI markers and stage sensitization model

25 volumetric and thickness markers were obtained from a total of 165 features using data from healthy controls, transition, early, intermediate and advanced AD cohorts. These include volumes and thickness of parahippocampal region and inferior temporal gyrus, entorhinal thickness in addition to hippocampal volumes. Most temporal lobe regions were identified as key markers predictive of HC and AD, excluding transverse temporal gyrus and banks of superior temporal sulcus. Fig. 3D shows extracted brain regions from MRI whose volumes/thickness are indicative of non-ageing but AD-related atrophy. List of extracted regions is given in Supplementary Table 1.

#### 3.2.1 Stage sensitization model

Logistic regression model built using these MRI markers demonstrated an accuracy of 82%, sensitivity and specificity of 88% and 82% with an AUC of 0.90 on the test dataset.

### 3.3 PET markers and amyloid proxy model

Along with global amyloid burden, mean SUVR in regions such as precuneus, lateral temporal regions, posterior cingulate, left entorhinal, left inferior temporal, right pericalcarine, right temporal pole were identified as some of the key differentiating regions. List of the 17 features extracted from both AV45 and PiB PET data is given in Supplementary Table 2.

#### 3.3.1 Amyloid proxy model

A logistic regression based amyloid proxy model demonstrated an accuracy of 90%, sensitivity and specificity of 87% and 90% with an AUC of 0.96 on test dataset.

### 3.4 Meta model

In this metamodel built with the three above independent models, MRI markers contributed the most to the classification model performance (Gain = 0.56), followed by PET features (Gain = 0.25) and Risk factors (Gain = 0.20). On test dataset, this model demonstrated an accuracy of 93%, sensitivity and specificity of 80% and 95% with an AUC of 0.95.

For comparison, a logistic regression model using risk factors and MRI features was also developed which demonstrated an accuracy of 91% and AUC of 0.93. Comparison of all individual and ensemble models, and their ROC are given in Table 2 and Fig. 4.

**Table 2.** Performance of individual and ensemble models.

|  | **Predisposition**<br>**model**<br>(Risk factors) | **Stage**<br>**sensitization**<br>**model** (MRI -<br>Atrophy) | **Amyloid**<br>**proxy model**<br>(PET tracer<br>uptake) | **Risk +**<br>**atrophy** | **Metamodel** |
| --- | --- | --- | --- | --- | --- |
| Accuracy | 69% | 82% | 90% | 91% | 93% |
| Sensitivity | 70% | 88% | 87% | 85% | 80% |
| Specificity | 68% | 82% | 90% | 92% | 95% |

**Fig. 4.**
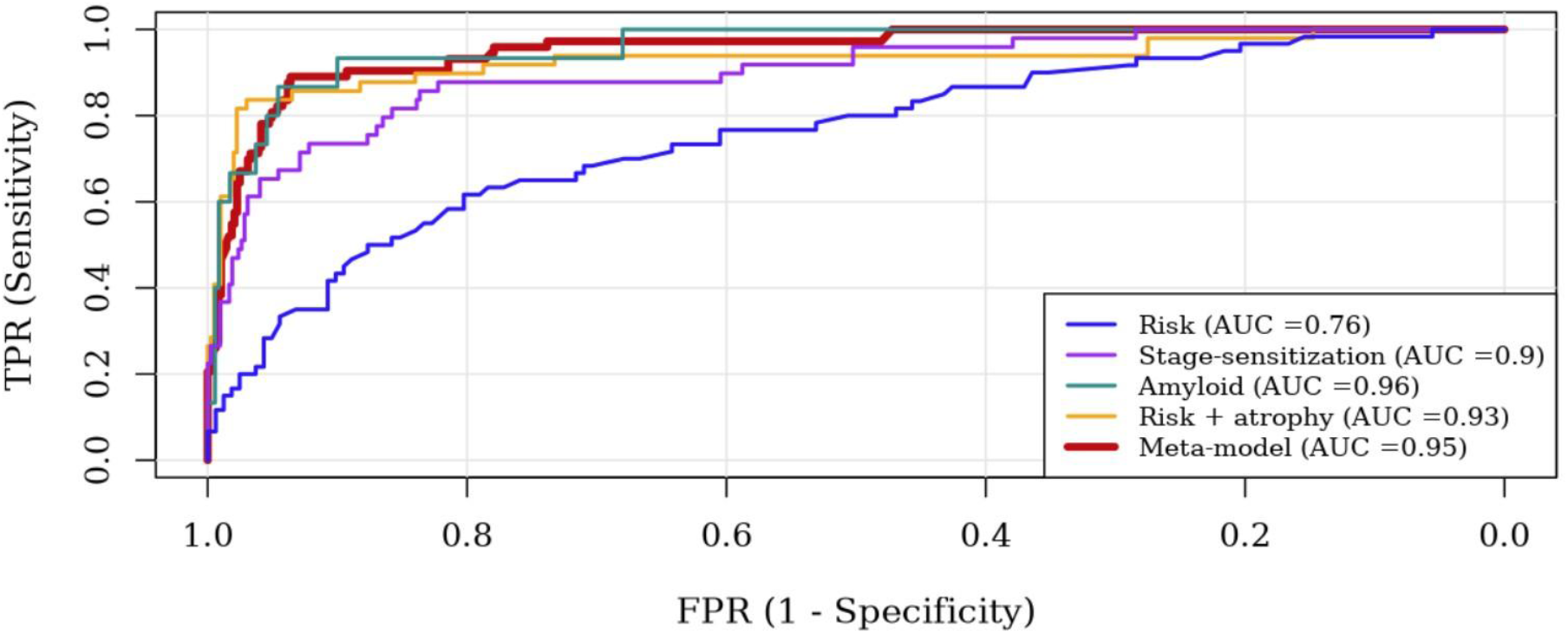
Receiver Operating Characteristic curves for models

## 4. Discussion

In this study, risk factors strongly associated with Alzheimer’s were extracted, MRI markers indicative of atrophy stages and PET markers indicative of amyloid deposition. An ensemble model predicting likelihood of an individual having AD has been built, which has showed improved performance and can estimate the likelihood of AD with reasonable accuracy. Even in the absence of PET data, model using risk factors and brain atrophy features together offer improved accuracy and specificity over individual models.

Consistent with the literature, strong associations between AD and age, APOE status and maternal dementia were found. Additionally, associations between AD and several medical conditions were also tested – specifically cardiovascular conditions (fibrillation, heart attack, heart failure, cardiac procedures - angioplasty, bypass or pacemaker procedures, other cardiovascular diseases), cerebrovascular conditions (stroke, transient ischemic attack, brain trauma, seizures, Parkinsons disease and parkinsonism disorders, cerebrovascular diseases and any other neurological conditions) and other conditions (alcohol depression, B12 deficiency, diabetes, hypercholesterolemia, hypertension, thyroid disease, fecal and urinary incontinence). Of the midlife risk factors noted in the 2024 Lancet commission report [22], depression, brain trauma, diabetes and hypertension were found to be associated with increased risk of AD in this study. However, no such similar associations were found with alcohol abuse and hypercholesterolemia which could also be attributed to the lack of completeness in the dataset used, which affects strength of analyses due to lower sample sizes.

The 3 different atrophy clusters within AD group obtained using PCA and hierarchical clustering correlated well with age and MMSE scores, with the lowest atrophy cluster (early AD group) being youngest and having highest mean MMSE scores and highest atrophy cluster (advanced AD group) being the oldest and having lowest mean MMSE scores. While this kind of exact delineation was not reflected in CDR scores, it is to be noted that majority have questionable or mild cognitive impairment, except for 7.4% of advanced AD group having moderate cognitive impairment. In CDR subscales, an average of 4% of individuals in transition AD group showed some mild or greater form of impairment, which rose to an average of 30% in early and intermediate AD groups. Interestingly, although the proportion of individuals with no or mild impairment was lower in the intermediate AD group, a higher proportion of individuals in this group also exhibited moderate levels of impairment across all subscales - except for Judgment and Problem-Solving subscale - compared to the early AD group. It’s in the advanced AD group where 61% of individuals had mild or greater impairment, with 80% affected in memory and 66% impaired in orientation, judgement and problem solving, home & hobbies subscales. Based on the analysis, personal care was the last of the CDR subscale domains affected in AD. Proportion of individuals with mild or greater level of impairment in AD groups is shown in Supplementary Figure. 1.

While hippocampal atrophy is a known marker, it alone is insufficient to monitor patients on the path of disease progression, especially as symptomatic changes are observed on CDR subscales. Our analyses revealed volumes of amygdala, fusiform and supramarginal gyrus as well as entorhinal cortex thickness in the right hemisphere and thickness of middle and inferior temporal gyrus in left hemisphere in addition to bilateral hippocampal volumes, as key discriminative factors in differentiating healthy controls and AD patients.

Similarly, in addition to global amyloid burden measured using mean cortical SUVR of precuneus, prefrontal cortex, gyrus rectus, and lateral temporal regions, regional amyloid patterns, seem to provide additional value for diagnosing AD. Tracer uptake in left inferior temporal region is found to play a significant role in tracking AD in both PiB based as well as AV45 PET imaging and common regions of interest between MRI and PET features were entorhinal cortex, inferior temporal region and temporal pole in temporal lobe, lingual and pericalcarine regions of occipital lobe and inferior parietal region. Amyloid proxy model built using these markers provided best overall performance, compared to other models even with sparse data. Further investigation using larger datasets could enable tracking amyloid deposition vis-à-vis atrophy levels and could provide holistic approach to understanding the progression of AD through in-vivo imaging marker tracking.

### 4.1 Limitations

A key limitation of this study is the incompleteness of the data, especially in risk factors and PET imaging analysis. Especially the predisposition model was found to be sensitive to sample sizes of medical history items. Therefore, to get exact risk estimates, larger and complete data is required. Additionally, PET imaging data was carried out with two different tracers and although tracer interaction is accounted for in this study, low sample sizes in AD groups limit the generalizability of the amyloid proxy model. As such, this study serves as proof-of-concept work on multimodal data integration for AD diagnosis using PET, MRI and risk factors and hence validation and reproducibility studies would be conducted by incorporating additional datasets to improve strength of the study. Similarly, the generalizability of the models will also be evaluated and updated through this inclusion of new data so that a clinically translatable and interpretable metamodel can be clinically deployed.

## Declarations

## Acknowledgements

Data analysis services contributing to this work were done on AD workbench which has been provided in-kind by the AD Data Initiative; McHugh CP, Clement MHS, Phatak M. AD Workbench: Transforming Alzheimer’s research with secure, global, and collaborative data sharing and analysis. Alzheimers Dement. 2025 May;21(5): e70278. doi: 10.1002/alz.70278. PMID: 40387289; PMCID: PMC12086970. https://www.alzheimersdata.org.

## Author contributions

Geetha Chilla has designed, executed and prepared this manuscript and its contents for this study.

## Ethics and Consent to Participate declarations

Not applicable. Data used in this study is de-identified and publicly available from OASIS-3 cohort, where all participants had provided informed consent at enrolment. As no new recruitment or direct interaction with human subjects was undertaken for this study, additional consent is not necessary.

## Consent to publish declaration

Not applicable

## Data availability

The data described in this study was provided and can be accessed on OASIS-3 under: Longitudinal Multimodal Neuroimaging: Principal Investigators: T. Benzinger, D. Marcus, J. Morris; NIH P30 AG066444, P50 AG00561, P30 NS09857781, P01 AG026276, P01 AG003991, R01 AG043434, UL1 TR000448, R01 EB009352. AV-45 doses were provided by Avid Radiopharmaceuticals, a wholly owned subsidiary of Eli Lilly. OASIS-3 data can be requested at https://sites.wustl.edu/oasisbrains/.

## Clinical Trial

Not applicable

## References

1. Bomasang-Layno E, Bronsther R (2021) Diagnosis and Treatment of Alzheimer’s Disease: An Update. Dela J Public Health 7:74. 10.32481/DJPH.2021.09.009

2. Braak H, Braak E (1991) Neuropathological stageing of Alzheimer-related changes. Acta Neuropathol 82:239–259. 10.1007/BF00308809

3. Braak H, Alafuzoff I, Arzberger T, et al (2006) Staging of Alzheimer disease-associated neurofibrillary pathology using paraffin sections and immunocytochemistry. Acta Neuropathol 112:389. 10.1007/S00401-006-0127-Z

4. Gold G, Bouras C, Kövari E, et al (2000) Clinical validity of Braak neuropathological staging in the oldest-old. Acta Neuropathol 99:579–582. 10.1007/S004010051163

5. Therriault J, Pascoal TA, Lussier FZ, et al (2022) Biomarker modeling of Alzheimer’s disease using PET-based Braak staging. Nature Aging 2022 2:6 2:526–535. 10.1038/s43587-022-00204-0

6. Ishii H, Meguro K, Yamaguchi S, et al (2006) Different MRI findings for normal elderly and very mild Alzheimer’s disease in a community: Implications for clinical practice: The Tajiri Project. Arch Gerontol Geriatr 42:59–71. 10.1016/J.ARCHGER.2005.06.002

7. Jack CR, Andrews JS, Beach TG, et al (2024) Revised criteria for diagnosis and staging of Alzheimer’s disease: Alzheimer’s Association Workgroup. Alzheimer’s & Dementia 20:5143–5169. 10.1002/ALZ.13859

8. Chapleau M, Iaccarino L, Soleimani-Meigooni D, Rabinovici GD (2022) The Role of Amyloid PET in Imaging Neurodegenerative Disorders: A Review. Journal of Nuclear Medicine 63:13S. 10.2967/JNUMED.121.263195

9. Ossenkoppele R, Pichet Binette A, Groot C, et al (2022) Amyloid and tau PET-positive cognitively unimpaired individuals are at high risk for future cognitive decline. Nature Medicine 2022 28:11 28:2381–2387. 10.1038/s41591-022-02049-x

10. Chen SD, Lu JY, Li HQ, et al (2021) Staging tau pathology with tau PET in Alzheimer’s disease: a longitudinal study. Translational Psychiatry 2021 11:1 11:1–12. 10.1038/s41398-021-01602-5

11. Maschio C, Ni R (2022) Amyloid and Tau Positron Emission Tomography Imaging in Alzheimer’s Disease and Other Tauopathies. Front Aging Neurosci 14:838034. 10.3389/FNAGI.2022.838034/PDF

12. Dallaire-Théroux C, Beheshti I, Potvin O, et al (2019) Braak neurofibrillary tangle staging prediction from in vivo MRI metrics. Alzheimer’s & Dementia: Diagnosis, Assessment & Disease Monitoring 11:599–609. 10.1016/j.dadm.2019.07.001

13. Whitwell JL, Josephs KA, Murray ME, et al (2008) MRI correlates of neurofibrillary tangle pathology at autopsy: A voxel-based morphometry study. Neurology 71:743–749. 10.1212/01.WNL.0000324924.91351.7D

14. Wisch JK, Petersen K, Millar PR, et al (2025) Cross-Sectional Comparison of Structural MRI Markers of Impairment in a Diverse Cohort of Older Adults. Hum Brain Mapp 46:. 10.1002/HBM.70133

15. Morrison C, Dadar M, Shafiee N, Collins DL (2023) The use of hippocampal grading as a biomarker for preclinical and prodromal Alzheimer’s disease. Hum Brain Mapp 44:3147. 10.1002/HBM.26269

16. Hebling Vieira B, Liem F, Dadi K, et al (2022) Predicting future cognitive decline from non-brain and multimodal brain imaging data in healthy and pathological aging. Neurobiol Aging 118:55. 10.1016/J.NEUROBIOLAGING.2022.06.008

17. Strittmatter WJ, Saunders AM, Schmechel D, et al (1993) Apolipoprotein E: high-avidity binding to beta-amyloid and increased frequency of type 4 allele in late-onset familial Alzheimer disease. Proc Natl Acad Sci U S A 90:1977. 10.1073/PNAS.90.5.1977

18. Corder EH, Saunders AM, Risch NJ, et al (1994) Protective effect of apolipoprotein E type 2 allele for late onset Alzheimer disease. Nature Genetics 1994 7:2 7:180–184. 10.1038/ng0694-180

19. Chen XR, Shao Y, Sadowski MJ (2021) Segmented Linear Mixed Model Analysis Reveals Association of the APOEε4 Allele with Faster Rate of Alzheimer’s Disease Dementia Progression. J Alzheimers Dis 82:921–937. 10.3233/JAD-210434

20. O’Neal MA (2023) Women and the risk of Alzheimer’s disease. Front Glob Womens Health 4:1324522. 10.3389/FGWH.2023.1324522/BIBTEX

21. Honea RA, Vidoni ED, Swerdlow RH, Burns JM (2012) Maternal family history is associated with Alzheimer’s disease biomarkers. J Alzheimers Dis 31:659–668. 10.3233/JAD-2012-120676

22. Livingston G, Huntley J, Liu KY, et al (2024) Dementia prevention, intervention, and care: 2024 report of the Lancet standing Commission. The Lancet 404:572–628. 10.1016/S0140-6736(24)01296-0

23. Shojaie M, Tabarestani S, Cabrerizo M, et al (2021) PET Imaging of Tau Pathology and Amyloid-β, and MRI for Alzheimer’s Disease Feature Fusion and Multimodal Classification. J Alzheimers Dis 84:1497. 10.3233/JAD-210064

24. Kim DH (2025) Longitudinal Analysis of Amyloid PET and Brain MRI for Predicting Conversion from Mild Cognitive Impairment to Alzheimer’s Disease: Findings from the ADNI Cohort. Tomography 11:37. 10.3390/TOMOGRAPHY11030037

25. Bao YW, Wang ZJ, Shea YF, et al (2024) Combined Quantitative amyloid-β PET and Structural MRI Features Improve Alzheimer’s Disease Classification in Random Forest Model - A Multicenter Study. Acad Radiol 31:5154–5163. 10.1016/j.acra.2024.06.040

26. Qiu S, Miller MI, Joshi PS, et al (2022) Multimodal deep learning for Alzheimer’s disease dementia assessment. Nature Communications 2022 13:1 13:1–17. 10.1038/s41467-022-31037-5

27. Tang Y, Xiong X, Tong G, et al (2024) Multimodal diagnosis model of Alzheimer’s disease based on improved Transformer. Biomed Eng Online 23:8. 10.1186/S12938-024-01204-4

28. Weinstein AM, Fang F, Chang CCH, et al (2024) Multimodal neuroimaging biomarkers and subtle cognitive decline in a population-based cohort without dementia. J Alzheimers Dis 103:570. 10.1177/13872877241303926

29. Aghdam MA, Bozdag S, Saeed F (2025) Machine-learning models for Alzheimer’s disease diagnosis using neuroimaging data: survey, reproducibility, and generalizability evaluation. Brain Informatics 2025 12:1 12:1–27. 10.1186/S40708-025-00252-3

30. LaMontagne PJ, Benzinger TLS, Morris JC, et al (2019) OASIS-3: Longitudinal Neuroimaging, Clinical, and Cognitive Dataset for Normal Aging and Alzheimer Disease. medRxiv 2019.12.13.19014902. 10.1101/2019.12.13.19014902

31. Raz N, Lindenberger U, Rodrigue KM, et al (2005) Regional brain changes in aging healthy adults: general trends, individual differences and modifiers. Cereb Cortex 15:1676–1689. 10.1093/CERCOR/BHI044

32. Tsagris M, Lagani V, Tsamardinos I (2018) Feature selection for high-dimensional temporal data. BMC Bioinformatics 19:1–14. 10.1186/S12859-018-2023-7/TABLES/3

33. Tsamardinos I, Aliferis CF, Statnikov A (2003) Time and sample efficient discovery of Markov blankets and direct causal relations. Proceedings of the ACM SIGKDD International Conference on Knowledge Discovery and Data Mining 673–678. 10.1145/956750.956838

34. Goldberg TE, Huey ED, Devanand DP (2020) Association of APOE e2 genotype with Alzheimer’s and non-Alzheimer’s neurodegenerative pathologies. Nature Communications 2020 11:1 11:1–8. 10.1038/s41467-020-18198-x

