## Supplementary for "Multimodal diagnosis of Alzheimer’s disease through causal imaging markers and risk factors"

Supplementary Table 1 – MRI markers

| MRI markers differentiating HC group with various atrophy stages of AD | MRI markers differentiating between AD stages |
|---|---|
| Right Hippocampus | Left entorhinal thickness |
| Left Hippocampus | Left inferiorparietal thickness |
| Left inferiortemporal thickness | Left inferiortemporal volume |
| Left middletemporal thickness | Left insula thickness |
| Right entorhinal thickness | Left isthmuscingulate thickness |
| Right fusiform volume | Left medialorbitofrontal volume |
| Right pericalcarine thickness | Left middletemporal thickness |
| Right supramarginal volume | Left superiorparietal volume |
| Right Amygdala | Left superiortemporal volume |
| | Right entorhinal thickness |
| | Right lingual thickness |
| | Right paracentral volume |
| | Right parahippocampal thickness |
| | Right parahippocampal volume |
| | Right precentral volume |
| | Right rostralmiddlefrontal volume |
| | Right temporalpole thickness |
| | Right Thalamus Proper |

Supplementary Table 2 – PET markers

| Markers from AV45 PET | Markers from PiB PET |
|---|---|
| Cortex - Left posterior cingulate cortex | Cortex - Left inferior temporal region |
| Cortex - Right posterior cingulate cortex | Cortex - Left inferior parietal region |
| Central Corpus Callosum | Right Putamen |
| Cortex - Right lateral occipital region | Left Accumbens |
| Cortex - Left inferior temporal region | Global Amyloid burden |
| WM - Left entorhinal region | Cortex - Right caudal middle frontal region |
| Cortex - Right pericalcarine | Right hemisphere Unsegmented White Matter |
| Cortex - Left lingual region | Cortex - Right temporal pole |
| | WM - Left Cuneus |
| | Left Caudate |

Supplementary Image 1

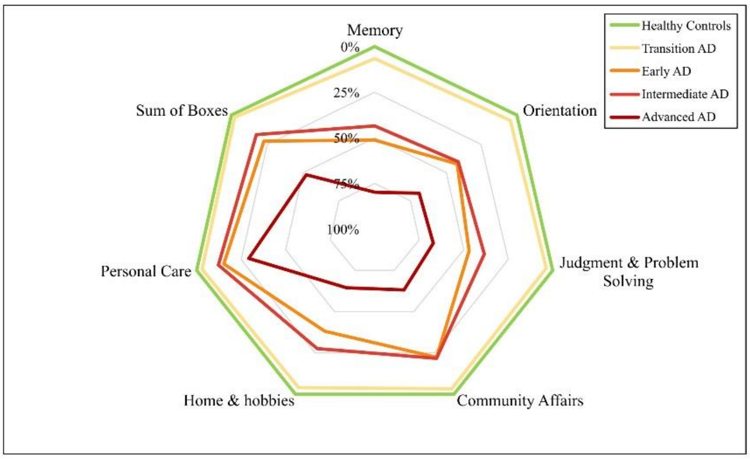


Supplementary figure 1 Proportion of AD patients with mild or greater impairment on CDR subscales by atrophy stage
